# Re-classification of archival Ovarian Carcinoma diagnostics using immunohistologic digital quantification and algorithmic prognosis

**DOI:** 10.1101/2020.08.05.20168849

**Authors:** Camelia D Vrabie, Mihnea I. Gangal, Marius Gangal

**Affiliations:** Senior Pathologist, Head of Pathology Department, “Sfântul loan” Clinical Emergency Hospital; Assist. Prof. at “Carol Davila” Medical University, Bucharest Romania; Surgical Gynecologist, “Notre Dame” Hospital, Montreal, QC, Ca; Assist. Prof. at “McGill” Medical University, Montreal, Canada; Research Pathologist, Medical Data Analytics Canada (MEDACS.ca), Rigaud, J0P1P0, QC, Canada

**Keywords:** archival ovarian carcinoma diagnostics, machine learning, random forest algorithm, immunohistochemistry digital quantification

## Abstract

Twenty years of research improved the classification of ovarian carcinoma, making the diagnostic relevant from a scientific and clinical perspective. Our research question was to find out if old studies are still pertinent under new diagnostic criteria and how we can use machine learning techniques for re-classification purposes.

The same main investigator re-classified 60 cases of ovarian carcinoma after 15 years, using 2014 WHO diagnostic criteria. Selected pathology data only (macro, micro information and immunohistochemistry images coming from a seven-stain panel) were provided for digital analysis. Biomarker images were digitalized and quantified using open source software and a validated methodology. 1080 attributes were classified using a random forest (open source) algorithm, using a supervised learning technique (the training dataset used 180 attributes). Human results were considered “ground truth” for the digital analysis.

The human analysis maintained the initial histopathologic diagnostic in 61.5% of cases. The digital prediction shows 80% accuracy and 73% precision when compared with human reclassified data. Based on results, we concluded that “recycling” of old studies is possible. Limitation of the study are the low number of cases analyzed, the total absence of clinical, treatment and prognostic data and a possible human criteria selection bias. Even if technical difficulties related to biomarker selection and histological analysis exist, digital investigation of existing, large archival registries is feasible, reliable and it can be done at a low cost.

## Introduction

A precise diagnostic of ovarian carcinoma (OC) is critical for the patient but sometimes is a challenge for the pathologist (^1^). Twenty years of research proved that OC is not a monolithic disease as it was presumed in the past. In 2004, Shih and Kurman proposed a dualistic model based on molecular and morphological features rather than only on histological appearance but it was difficult to apply outside research (^2^). Over time, molecular genetics, biomarker studies and clinical experience crystallized the actual concept of OC as a heterogenic disease. Five sub-classes were proposed, each originated from a distinct precursor cell: high-grade serous (HGSC), low-grade serous carcinoma (LGSC), endometroid (EC), mucinous (MC) and clear cell carcinoma (CCC). Each sub-class has a distinct natural history, distinct pathogenic pathway, different response to treatment and different prognostic (^3^). The new classification concept (WHO 2014) proved to have both clinical and diagnostic benefits and was rapidly adopted (^4^). From a pathologist’s perspective, the new classification strategy showed robust validity and allowed precise, reproducible diagnostics with reduced inter-observer variability (^5^).

Because possible observational variability and subjectivity in morphological data interpretation (^6^) other histologic techniques are necessary for increasing diagnostic precision. Immunohistochemistry (IHC), is able to detect specific tissues biomarkers and is used routinely in order to bring objectivity to the morphologic interpretation. It is widely adopted by surgical pathology laboratories but has several well-known limitations: staining interpretation is semiquantitative and not entirely standardized, fact that makes the method time consuming and subjective as well. Some IHC stains lack tumor specificity or react to proteins in normal or benign tumoral tissue. A way to circumvent these limits is by using IHC diagnostic panels. The use of multiple IHC will increase diagnostic precision but results interpretation may become complex and also diagnostic costs can soar. Concomitant interpretation of histologic and IHC panel data can make diagnostic interpretation even more challenging.

A way to improve the classification equation is by using artificial intelligence for the analytic process, a subject under investigation (^7^,^8^). Machine learning (ML), a part of Artificial Intelligence scientific domain, is a way a machine will learn from data itself rather from programming how to connect, sort and analyze information using predetermined algorithms (^9^). A machine can be instructed about possible results (supervised learning) or not (unsupervised learning) and will use classification, regression, clustering and association algorithms for data interrogation. For interpretation of complex data, ML algorithms allows fast, reliable and reproducible results. Supervised ML is probably close to the way humans are approaching pathology diagnostic: based on characteristics of a given set of data (“training set”), the machine will sort and connect information found in a “test dataset” and will generate predictions.

As the landscape of OC diagnostic significantly changed in the last 20 years, a legitimate question arises: are old studies still useful? Several authors re-visited old OC research datasets in order to investigate how recent diagnostic criteria changed the tumor classification paradigm over time and if old data can be of use in future research. Main barriers for old data use were classification changes and a constant deprecation of some IHC panel stains. Results were divergent. Kommoss (^10^) considered old OC studies of low value, with a high degree of inter and intra-observational variability. Peres (^11^) and Köbel (^12^) considered that historic histotype based datasets are still useful if data is re-analyzed under specific constrains.

The objective of our study was to evaluate how digital technology (digital analysis of microscopic images and the use of a machine learning classification algorithm) is able to re-classify an OC diagnostics dataset created in 2005 (^13^) based only on pathology information.

## Material and method

An archival dataset of 60 diagnosis of ovarian carcinoma (2005) was reclassified by the main investigator (CV) based on 2014 WHO criteria (table 1). Seven IHC markers were also selected from the original ten IHC panel stains, based on microscopic slides availability, literature and investigator’s experience (table 2). Digital images were taken using a standard microscope. The investigator established a Region of Interest (ROI) for each IHC stain image based on an area of maximal staining. All data was included in a “diagnostic” dataset (60 instances, 18 attributes). A “training” dataset was created based on 10 unequivocal OC histotypes.

**Table 1.** Morphologic attributes used in dataset re-classification (Ma: macroscopic observation, Mi: microscopic observation)

| Attribute | Measure | Variable | Type |
| --- | --- | --- | --- |
| Age | Years | Numeric | Ma |
| Tumor dimension | Cm | Numeric | Ma |
| Solidity | Low, Medium, High | Nominal | Ma |
| Necrosis | Yes, No | Nominal | Ma |
| Bilaterally | Yes, No | Nominal | Ma |
| Pattern | Micro-papillary, Papillary, Tubular, Cystic, Solid | Nominal | Mi |
| Fibrosis | Absent, Low, Medium, High | Nominal | Mi |
| Psammoma Bodies | Yes, No | Nominal | Mi |
| Large nucleus | Yes, No | Nominal | Mi |
| Mitosis | Yes, No | Nominal | Mi |
| Clear cells | Yes, No | Nominal | Mi |

**Table 2.** Immunologic markers used in dataset reclassification

| IHC attribute | Measurement | Type | Cut-off |
| --- | --- | --- | --- |
| Estrogen Receptor | Index | Numeric | No |
| Progesterone Receptor | Index | Numeric | No |
| CA-125 | Index | Numeric | No |
| p53 | Index | Numeric | No |
| Ki-67 | Index | Numeric | No |
| Her-2 | Yes, No | Nominal | 1% |
| CEA | Yes, No | Nominal | 15% |

The digital re-classification included digital data analysis and algorithm prediction. In the digital data analysis, each ROI from IHC image was digitally quantified with the help of an open label software (^14^). An optical density index (ODI) was determined based on ROI histogram (^15^). Using a dedicated plugin (IHC profiler), the total stained surface was measured in terms of pixel intensity of the visualization stain, (fig 1, 2) (^16^). An IHC stain index (SI) was calculated for each of five IHC markers (positive multiplied by ODI). For the other two markers, a low cut-off intensity was established and the presence of marker was defined as present or absent (table 2)

**Fig 1.**
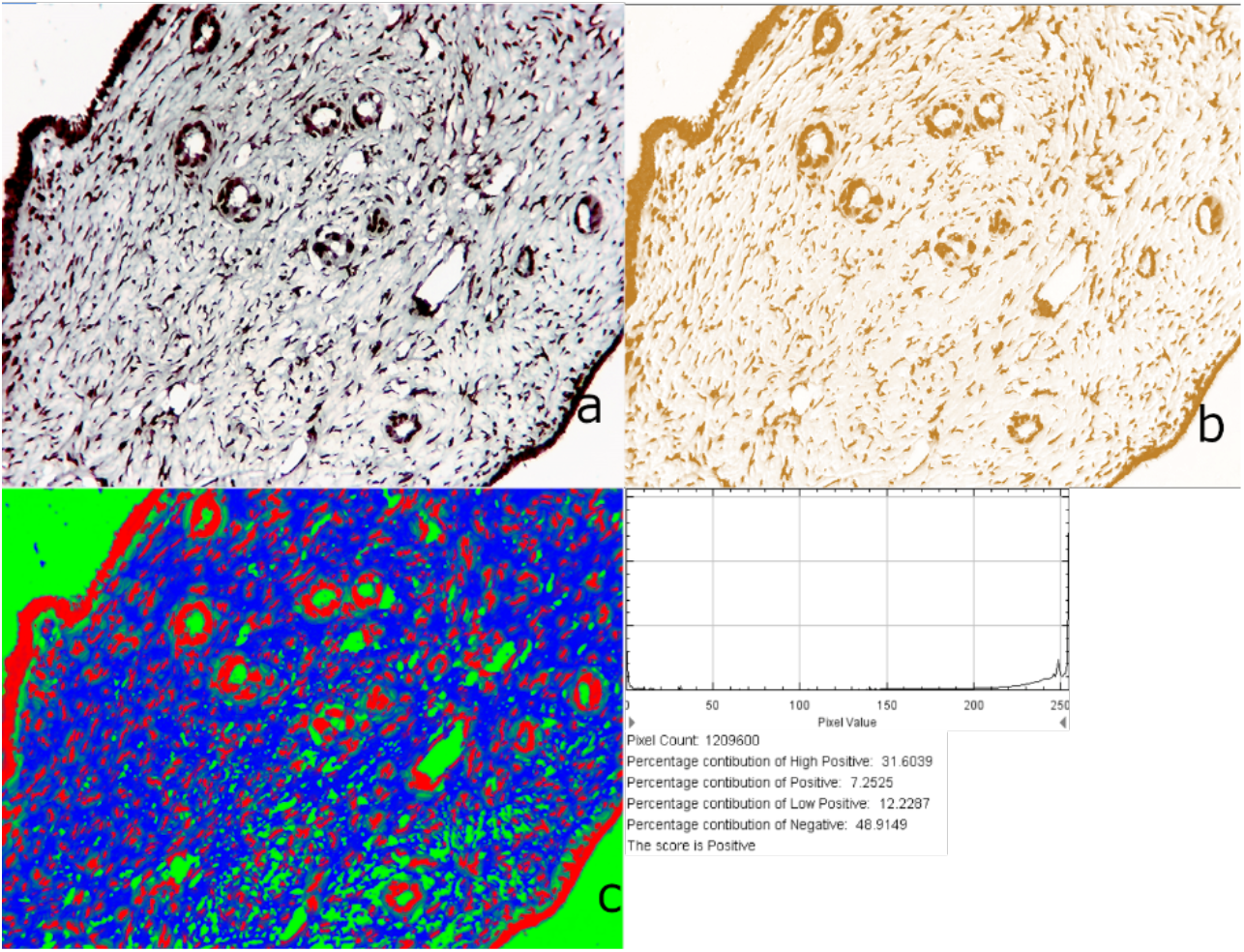
Ki67 quantification. a) Original image (ROI) (x200). b) deconvoluted, 8-bit image. c) probability analysis. Histogram and log showing 51.1% of image stained positive. ODI=0.264. SI=8.6

**Fig 2.**
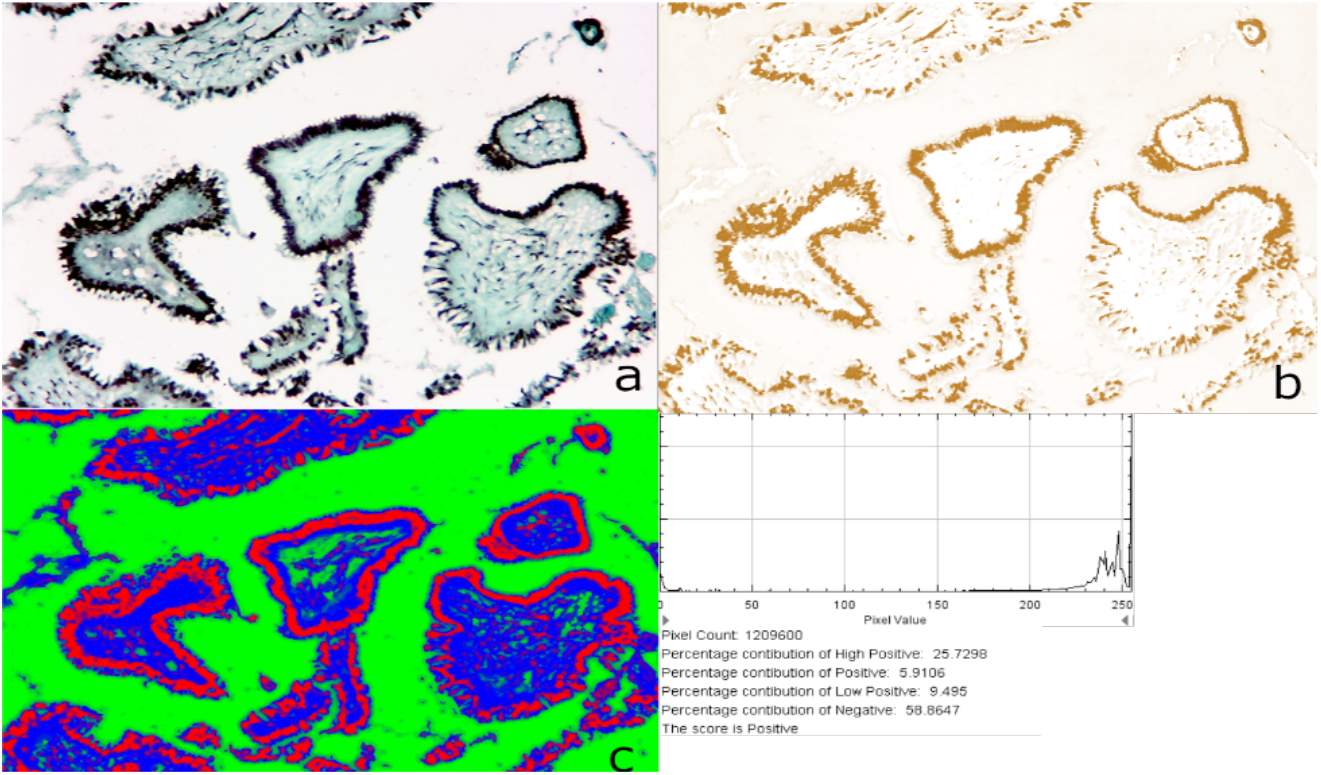
p53 quantification. a) Original image (ROI) (x200). b) deconvoluted, 8-bit image. c) probability analysis. Histogram and log showing 41.2% of image stained positive. ODI=0.202. SI=8.3

For ML classification we used a random forest supervised trained algorithm. We opted for an open source machine learning toolkit (WEKA) software (^17^) solution that is widely accepted in bioinformatics, has a good graphical interface and requires no programming skills (^18^).

Human reclassification results were considered “ground truth” for the digital investigation. The accuracy and precision (^19^) of algorithm prognosis was judged against the “ground truth”. Algorithm results were considered “true” when classification was concordant. Diagnostics were considered “positive” for serous carcinoma (the most clinically significant). Any other diagnostic was considered “negative”.

No clinical, treatment or prognostic information was available for the digital re-classification. The initial pathological methodology for sampling and staining of the microscopic slides (for both morphology and Immunohistologic characterization) was already described in the original publication.

### Statistics

All data was stored in a standard spreadsheet. Numerical data was analyzed using MedCalc Statistical software (version 19.0.7, MedCalc Software bvba, Ostend, Belgium). Basic statistic results were expressed as mean ± standard deviation. Because of the low number of cases in some categories, differences between means were not tested for significance. More info concerning the random forest classification algorithm can be found here (^20^)

### Ethics

Patients provided written, informed consent before the initial treatment, specifically agreeing with the processing and analyze of the pathology samples. As the study used only archived, published data, a new IRB approval was waived by the hospital IRB committee.

## Results

60 OC archived diagnostics were reviewed and reclassified by the same pathologist, after 15 years. The initial cohort contained 45 serous, 12 mucinous and 3 borderline carcinomas. The human re-classification produced 37 HGSC, 2 LGSC, 11 EC, 6 MC and 4 CCC. The algorithm prognosis generated 35 HGSC, 2 LGSC, 4 EC, 15 MC and 4 CCC diagnostics. The original Mucoid diagnostic was maintained in 25% of cases by human and 50% by machine. The Serous carcinoma diagnostic was maintained in 75% of cases by human and in 71% of cases by machine.

In 48 cases (table 3), the diagnostic was concordant: 36 cases were “true positive” (34 HGSC, 2 LGSC), 12 were “true negative” (2 EC, 4 CCC and 6 MC). In 12 cases the diagnostic was divergent (table 4): 8 cases were “false positive” (EC classified as MC; No MC case was classified as EC). 4 cases were “false negative” (2 HGSC as EC, 1 HGSC as MC, 1 EC classified as HGSC) (fig 2).

**Table 3.**
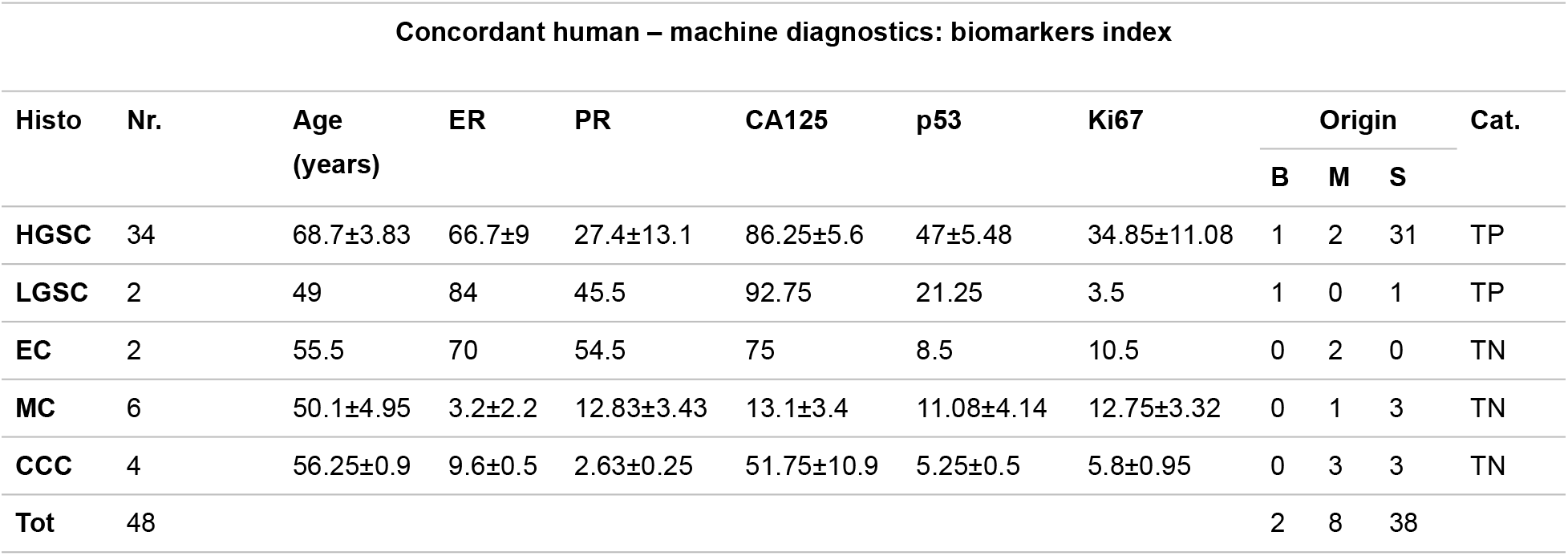
Concordant Diagnostics. Age (mean ± standard deviation). Calculated Index for selected biomarkers (ER, PR, CA125, p53 and Ki67): mean ± standard deviation. Origin: initial (2005) OC classification. TP: true (concordant) positive (serous). TN: true (concordant) negative (non-serous)

| Concordant human – machine diagnostics: biomarkers index |  |  |  |  |  |  |  |  |  |  |  |
| --- | --- | --- | --- | --- | --- | --- | --- | --- | --- | --- | --- |
| Histo | Nr. | Age<br>(years) | ER | PR | CA125 | p53 | Ki67 | Origin |  |  | Cat. |
|  |  |  |  |  |  |  |  | B | M | S |  |
| HGSC | 34 | 68.7±3.83 | 66.7±9 | 27.4±13.1 | 86.25±5.6 | 47±5.48 | 34.85±11.08 | 1 | 2 | 31 | TP |
| LGSC | 2 | 49 | 84 | 45.5 | 92.75 | 21.25 | 3.5 | 1 | 0 | 1 | TP |
| EC | 2 | 55.5 | 70 | 54.5 | 75 | 8.5 | 10.5 | 0 | 2 | 0 | TN |
| MC | 6 | 50.1±4.95 | 3.2±2.2 | 12.83±3.43 | 13.1±3.4 | 11.08±4.14 | 12.75±3.32 | 0 | 1 | 3 | TN |
| CCC | 4 | 56.25±0.9 | 9.6±0.5 | 2.63±0.25 | 51.75±10.9 | 5.25±0.5 | 5.8±0.95 | 0 | 3 | 3 | TN |
| Tot | 48 |  |  |  |  |  |  | 2 | 8 | 38 |  |

**Table 4.** Non-concordant Diagnostics. Age (mean ± standard deviation). Calculated Index for selected biomarkers (ER, PR, CA125, p53 and Ki67): mean ± standard deviation. Origin: initial (2005) OC classification. FP: false (non-concordant) positive (non-serous). FN: false (non-concordant) negative (serous)

| Non-concordant human machine Diagnostics |  |  |  |  |  |  |  |  |  |  |  |  |
| --- | --- | --- | --- | --- | --- | --- | --- | --- | --- | --- | --- | --- |
| Histo |  | Nr | Age | ER | PR | CA125 | p53 | Ki67 | Origin |  |  | Cat. |
| Human | Machine |  |  |  |  |  |  |  | B | M | S |  |
| HGSC | EC/MC | 3 | 69.67±1.52 | 54.33±8.5 | 17.16±4.75 | 89.83±2.25 | 46.16±1.15 | 14.5±2.29 | 0 | 1 | 2 | FN |
| EC | HGS | 1 | 57 | 71 | 8.5 | 80 | 72.5 | 12 | 0 | 0 | 1 | FN |
| EC | MC | 8 | 55.37±1.5 | 25.5±10.6 | 7.25±1.28 | 72.5±11.45 | 16.38±4.04 | 10.81±1.25 | 1 | 3 | 4 | FP |
| Tot |  | 12 |  |  |  |  |  |  | 1 | 4 | 7 |  |

**Fig 2.**
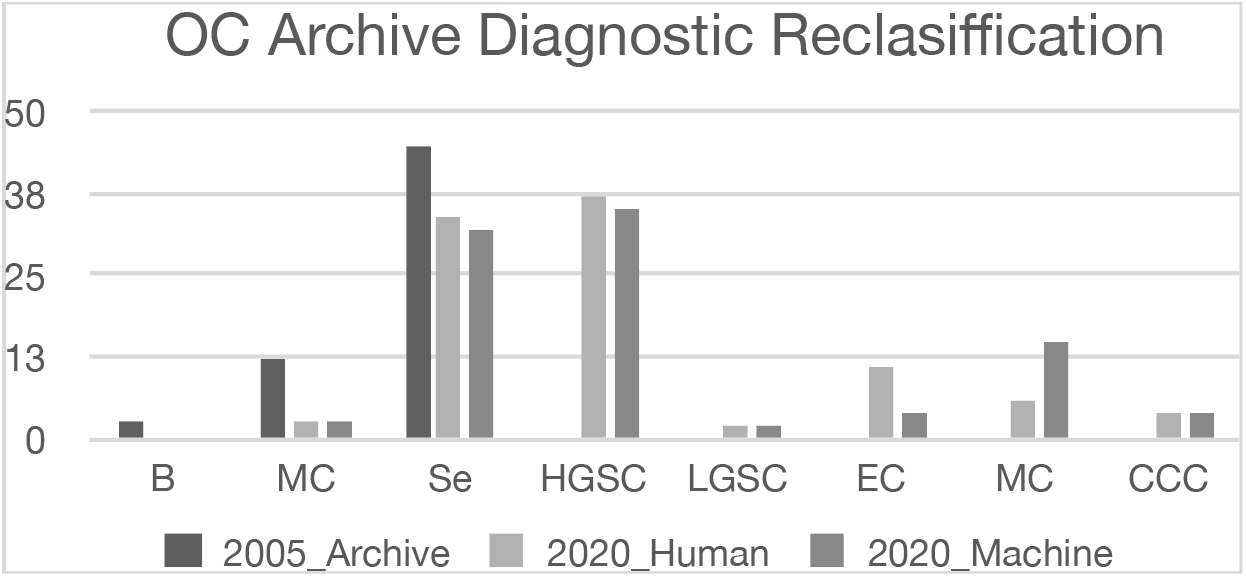
Reclassification of OC diagnostics based on WHO 2014 histotype criteria (Human – main investigator; Machine – random forest classifier)

## Discussions

Ovarian cancer represents more than 30% of all cancer cases of the female genital tract. 90% of them are carcinomas, originated from different precursor ovarian epithelium cells. Following new genomic, histologic and clinical data, the OC classification criteria changed in time, fact that improved clinical response to therapy and disease prognostic. Recent classification (WHO 2014) still maintain histologic differentiation as the main classification factor, fact that create place for inter and intra-observational variability. To improve diagnostic precision, large IHC panels can be used (up to 22 biomarkers) but results interpretation can be challenging. A possible way to interpret large datasets in which morphology, IHC, genomics, clinical and treatment information have to be analyzed together is provided by digital technology. ML algorithms can sort and interpret large datasets in a fast, reliable and reproducible way. ML algorithms can combine digital pathology data with any socio-demographic, macro and microscopic data for predicting diagnostics based on new diagnostic classification criteria. Good classification results were already published.

Our study question was to investigate if an old OC study (2005) may still have scientific validity, in the context of changing both of disease classification criteria and IHC panel structure. Our presumption was that digital technology will allow “recycling” of old studies possible, saving time and resources, both important in basic research.

For reclassification we used only pathology data (macroscopic, microscopic data and IHC images) selected by the main (initial) investigator. The dataset (60 instances) was interrogated using a random forest algorithm. The “training” dataset included 10 unquestionable diagnosed OC subtypes (two per subtype) and was provided by the main investigator as well.

Under 2014 OC classification criteria, the human investigator confirmed 37 (61.5%) from initial diagnostics (representing 25% of mucinous and 75% of serous initial diagnostics). The random forest algorithm confirmed 35 (58%) of the initial diagnostics (50% mucinous and 71% serous).

The algorithm classification performed well with 80% accuracy and 73% precision (90% sensitivity and 60% specificity) in reclassification of serous carcinoma. The most challenging diagnostic for the algorithm was the EC-MC classification, and that can allow an explanation for the low sensitivity (8 cases – 13%). The relative high number of errors (8) was probably generated by the absence of a specific stain in the used IHC panel to allow differentiation between non-serous carcinoma. The most serious error (4 cases – 6.67%) was the classification of HGSC tumors as EC or MC. One essential morphology criterion used for serous tumors differentiation was using nominal (large nucleus – yes/no) and not a numeric value and that created the diagnostic confusion. Digital segmentation and nuclear measurement will certainly improve the classification. On the other hand, even when narrative data was used, a precise differentiation of some histotypes (LGSC and CCC) was still possible.

Other limitations of the study were the absence of any clinical, staging or prognostic information and the low number of cases included. In order to reduce overfitting of the machine analysis, even important pathology pieces of information were blinded. Technical difficulties of IHC data capture on archived slides have to be also emphasized. Re-staining of the tissue (when possible) and the use of tissue microarrays for IHC analysis are solutions for improving archival analysis but the tissue can be scarce and supplementary analysis will certainly increase the costs of re-classification.

With an 80% accuracy, algorithm OC classification can be used in research. “Recycling” old research data is possible, and with the help of digital technologies, research costs can be reduced significantly. The process may be of interest for some research centres as the costs increase and availability of tissue samples decrease. It will also give researchers an idea about the disease trends in time and a way to homogenize diagnostics coming from different research registries.

## Data Availability

all data was already in public domain, reclassification machine learning study

## Author Statement

CV and MDG conceived the idea, CV performed the histologic evaluation, MDG performed the digital image acquisition, digital analysis and first draft conception. MIG corrected and finalized the paper. All authors reviewed the final form and agreed on publication final version.

## References

1 Prat J., New Insights into ovarian cancer pathology, Annals of Oncology 23 (Supplement 10): x111–x117, (2012) doi: 10.1093/annonc/mds300

2 Shih M, Kurman RJ. Ovarian tumorigenesis: a proposed model based on morphological and molecular genetic analysis. Am J Pathol. (2004) 164:1511–8. doi: 10.1016/S0002-9440(10)63708-X)HGSCCC

3 Köbel M, Kalloger SE, et al (2008) Ovarian carcinoma subtypes are different diseases: implications for biomarker studies. PLoS Med 5(12): doi: 10.1371/journal.pmed.0050232

4 Kurman RJ, Carcangiu ML, Herrington CS, Young RH (eds) (2014) WHO Classification of Tumours of Female Reproductive Organs. 4th edn. IARC Press: Lyon, France

5 Köbel M, Bak J, et al (2014) Ovarian carcinoma histotype determination is highly reproducible, and is improved through the use of immunohistochemistry. Histopathology 64(7): 1004–1013, doi: 10.1111/his.12349

6 Hernandez E, Bhagavan BS, et al Interobserver variability in the interpretation of epithelial ovarian cancer., Gynecol. Oncol 17 (1984) 117–23. http://www.ncbi.nlm.nih.gov/pubmed/6693048

7 Kalloger SE., Köbel M, Calculator for ovarian carcinoma subtype prediction Modern Pathology (2011) 24, 512–521, doi:10.1038/modpathol.2010.215

8 Wu M., Yan C., Automatic classification of ovarian cancer types from cytological images using deep convolutional neural networks, Bioscience Reports (2018) 38 0289, Doi: 10.1042/BSR20180289

9 www.IBM.com/analytics, accessed on May 19, 2020

10 Kommoss S, Gilks CB, et al, Ovarian carcinoma diagnosis: the clinical impact of 15 years of change, Br. J. Cancer 115 (2016) 1–7. 10.1038/bjc.2016.273

11 Peres L., Cushing-Haugen K.L., et al, Histotypes classification of ovarian carcinoman: a comparison of approaches, Gynecol Oncol. 2018 October; 151(1): 53–60. doi:10.1016/j.ygyno.2018.08.016

12 Köbel M., Kalloger SE., Biomarker-Based Ovarian Carcinoma Typing: A Histologic Investigation in the Ovarian Tumor Tissue Analysis Consortium, Cancer Epidemiol Biomarkers Prev; 2013, 22(10); 1677–86 2013, DOI: 10.1158/1055-9965.EPI-13-0391

13 Vrabie CD, Petrescu A, et al, Clinical factors and biomarkers in ovarian tumors development, 2008, Rom J Morphol Embryol. 2008;49(3):327–38

14 Schindelin J, Arganda-Carreras I, et al Fiji: an open-source platform for biological-image analysis, Nature methods 2012 9(7): 676–682; doi: 10.1038/nmeth.2019

15 Ruifrok AC, Johnston DA (2001) Quantification of histochemical staining by colour deconvolution. Anal Quant Cytol Histol 23: 291–299

16 Varghese F, Bukhari AB, Malhotra R, De A (2014) IHC Profiler: An Open Source Plugin for the Quantitative Evaluation and Automated Scoring of Immunohistochemistry Images of Human Tissue Samples. PLoS ONE 9(5): e96801. doi:10.1371/journal.pone.0096801

17 Eibe Frank, Mark A. Hall, and Ian H. Witten (2016). The WEKA Workbench. Online Appendix for “Data Mining: Practical Machine Learning Tools and Techniques”, Morgan Kaufmann, Fourth Edition, 2016

18 Frank E., Hall M., Data mining in bioinformatics Bioinformatics, Volume 20, Issue 15, 12 October 2004, Pages 2479–2481, https://doi.org/10.1093/bioinformatics/bth261

19 Eusebi P., Diagnostic Accuracy Measures, Cerebrovasc Dis 2013;36:267–272 DOI: 10.1159/000353863

20 Amrhen M., Muall F., The Random Forest Classifier in WEKA: Discussion and New Developments for Imbalanced Data, (2019) arXiv:submit/2529128

